# Global Adoption of openEHR Clinical Data Repositories: A Vendor and Community Survey

**DOI:** 10.64898/2026.08.27.26361529

**Authors:** Severin Kohler, Falk Meyer-Eschenbach, Xabier Michelena, Nicolas Frey, Michael Marschollek, Roland Eils

## Abstract

The openEHR standard provides an open, vendor-neutral architecture for clinical data repositories (CDRs), yet its real-world deployment has not been systematically documented. We conducted a dual-perspective survey combining a vendor survey of openEHR CDR providers with a community survey of openEHR practitioners. Eleven vendor organisations reported deployments across 21 countries and over 100 institutions and health regions. A complementary community survey (n=29, 17 countries) provided context on regulatory environments, adoption drivers, and barriers. Combined, the surveys cover 28 countries, 25 of them with a reported openEHR CDR deployment. Three findings emerge: openEHR has achieved national-scale presence through two distinct channels. Through vendor-market convergence, openEHR-based systems cover the majority of regional health authorities without a national mandate, including 19 of 21 Swedish regions, 3 of 4 Norwegian health regions, and 16 of 21 Finnish wellbeing services counties. Through national health record adoption, governments have built or procured national systems on openEHR as their technical foundation, including Ireland, Malta, Greece, Slovenia, and, by community report, Iran. Across Europe, this constitutes an openEHR-based interoperability infrastructure already in place across multiple EU member states. We identified no country in which openEHR is named in binding national regulation, creating structural fragility and an unrealised opportunity for alignment with the European Health Data Space (EHDS). Second, 61% of deployments serve primary use only, and 12% support both primary and secondary use. Third, lack of openEHR-specific knowledge is the most consistent adoption barrier across all geographies and deployment tiers. Adoption is driven by practitioner need and innovation, not by regulatory mandate.

## 1 INTRODUCTION

Semantic interoperability in healthcare information systems remains a central challenge in medical informatics(1). As clinical data volumes grow, the ability to store, exchange, and reuse data across institutional and national boundaries has become a prerequisite for both patient care and research(2). Several open standards have emerged to address different aspects of this challenge. HL7 FHIR targets data exchange(3, 4), the OMOP Common Data Model targets observational research(5, 6), and openEHR targets persistent clinical data storage and modelling(7). These standards have been proposed to serve complementary rather than competing roles(8, 9).

The openEHR standard employs a two-level modelling approach, separating a stable reference model from domain-specific clinical content defined through archetypes and templates(10, 11). This architecture allows clinical domain experts to define data structures independently from software developers(12), supporting semantic interoperability and future-proof data persistence. Archetypes are reusable across languages and clinical contexts(13), and the openEHR Query Language (AQL) enables structured retrieval from archetype-based repositories(14, 15).

Technical feasibility of openEHR has been demonstrated across multiple clinical domains, including decision support(16), natural language processing(17), and research infrastructure such as the German HiGHmed consortium(18). Integration pathways to OMOP(19, 20) and HL7 FHIR(21) have been developed. Despite over two decades of development, the actual deployment landscape of openEHR clinical data repositories (CDRs) has not been systematically documented in the peer-reviewed literature.

Frade et al. identified 21 openEHR projects in a 2013 survey(22). Delussu et al. surveyed 19 openEHR CDR vendors and grouped the responding systems into four functional clusters from a technical perspective(23). Neither study assessed where and how CDRs are deployed in production, what institutions use them, or whether they serve primary clinical use, secondary research use, or both. Definitions of these terms vary in the literature. Throughout this article, primary use denotes the use of clinical data for the direct care of the individual patient, while secondary use denotes use for purposes other than those for which the data were originally collected, such as research, quality improvement, and public health(24, 25). This gap matters because the value proposition of openEHR depends on its ability to support the learning health system vision, in which routine clinical data is reused for research, quality improvement, and public health(26, 27, 24).

This study conducts the first vendor-based survey dedicated to mapping global openEHR CDR adoption, complemented by a community survey on regulatory environments, adoption drivers, and barriers. Specific aims were to map the geographic distribution of openEHR CDR deployments, to characterise deployment depth by national, regional, or institutional scope, to assess the primary-secondary use balance, and to identify adoption patterns from the practitioner perspective.

## 2 METHODS

### 2.1 Study design and participants

We conducted two independent cross-sectional surveys: a vendor-based survey of openEHR CDR providers and a community survey of openEHR practitioners. The two surveys targeted distinct, non-overlapping populations. The vendor survey addressed organisations that develop or deploy CDR products, while the community survey addressed individual practitioners, implementers, and policy stakeholders. The surveys were linked retrospectively at the country level to compare vendor-reported deployment facts with community-reported perceptions.

Vendor survey respondents were identified through the openEHR Foundation vendor directory, the openEHR community network, and prior publications on openEHR implementations. Sixteen organisations were contacted. The community survey was distributed via openEHR Foundation mailing lists and networks. The vendor survey collection period (October 2025 to February 2026) overlapped with the community survey (October 2025 to March 2026). All results reflect the deployment status reported in these periods, and no survey data collected after March 2026 were added. Reported entries were corrected only where follow-up established them to be inaccurate (see Discussion). Respondents reported on deployments they themselves develop or operate. We therefore treated their responses as good-faith expert accounts and took the reported deployment facts at face value, while not independently auditing them. Given the self-selected respondent pool, community survey data are used mainly to contextualise vendor findings in the Discussion.

### 2.2 Survey instruments and data processing

The vendor survey comprised eight questions collected via Google Forms, covering vendor country, company name, all institutions where the CDR is deployed, use type per deployment (primary only, secondary only, or dual use), and whether the CDR serves as an outpatient or hospital information system. For questions 3 through 8, respondents listed institutions by name and country. The community survey comprised 54 items covering adoption maturity (Likert scales), regulatory environment, adoption drivers and barriers, use of complementary standards including HL7 FHIR, and expected adoption trajectory. Both survey instruments are reproduced in full in the Supplementary Material. Vendor identities were anonymised and results are reported by country rather than by vendor name.

Vendor survey responses were received in two formats: ten respondents provided free-text answers, while one respondent submitted a structured table with one row per institution and binary use-type indicators. Free-text responses were manually curated to extract institution records, normalise country names, and resolve ambiguities. Aggregate figures (e.g., “170 institutions in Australia”) were recorded as single entries with counts preserved. All records were unified into a common schema with fields for vendor, institution, country, primary use, secondary use, and deployment scope. Deployment scope was classified as *national* (covering a national health system or majority of a country’s regions), *regional* (covering one or more administrative health regions), or *institutional* (individual hospitals or organisations), based on respondent descriptions and publicly available health system information. National scale was assigned where a respondent reported coverage of a majority of a country’s regions with an explicit numerator and denominator, or where a national government or ministry had procured a national health record. Deployments reported only as an institution count, without a coverage fraction, were not classified as national. Where a response was ambiguous or reported only as an aggregate, we sought clarification from the respondent by direct follow-up where feasible, in writing or in conversation. Because these exchanges were unstructured, they are not reproduced in the Supplementary Material. The clarifications obtained were recorded and analysed in the same anonymised, country-level form as the survey responses.

### 2.3 Analysis

Quantitative analysis comprised descriptive statistics of deployment counts, geographic distribution, vendor characteristics, and use-type distribution, stratified by deployment scope. Free-text responses from both surveys were analysed by inductive thematic summary. Two authors (SK, FME) independently reviewed qualitative data and identified themes, and independently assigned deployment scope to each country. Disagreements in both cases were resolved through discussion and consensus. Community survey regulatory facts and barrier themes were used to contextualise vendor-reported patterns. Regulatory status was assessed from community survey responses and from publicly available national policy and legislative documents for countries with national-scale deployments. No systematic review of national legislation was undertaken. Per-country satisfaction and Likert-scale scores were not used as evidence given the small and self-selected sample. Geographic visualisations were generated using geopandas and matplotlib in Python. Claude Code (Anthropic; models Sonnet 4.6 to Opus 5.5, April to November 2026) assisted with code for data processing and figures, structuring of the survey findings, and language editing of the manuscript and supplement. The authors reviewed and verified all content and take full responsibility for the work. The Clinical Research Ethics Committee of IDIAP Jordi Gol (Barcelona, Spain) determined that ethics approval was not required, as the study used anonymised data only and involved no identifiable health information or human subjects.

## 3 RESULTS

Eleven of 16 contacted organisations responded to the vendor survey (69%), headquartered across nine countries spanning Europe, Asia, and Oceania. Respondents described deployments in different terms: some listed individual institutions, others reported regional coverage, national health records, or aggregate institution counts. openEHR CDR deployments were reported in 21 countries across four continents. The community survey received 29 responses from 17 countries. Fig. 1 additionally marks four countries reported only in the community survey, namely Iran, Japan, Thailand, and the United States, for 25 in total. Of these, Iran was reported as a government-built national-scale deployment by a ministry-level respondent. Two additional countries were represented only in the community survey without a reported openEHR deployment, and a vendor-reported national record in Jamaica was excluded because it did not reach production (see Discussion), for 28 countries covered by the two surveys combined. The geographic distribution is strongly concentrated in Europe. Non-European deployments were vendor-reported in Australia, China, India, Indonesia, and Nigeria. Deployment scope varied substantially by country and is detailed in Table 1.

**Table 1.** Vendor-reported openEHR CDR adoption by country. Countries are grouped by deployment scope as classified by the authors based on vendor descriptions. All data are vendor self-reported and were not independently verified.

| Country | Sources | Primary | Secondary | Reported scale |
|---|---|---|---|---|
| *National adoption* |  |  |  |  |
| Finland | 2 | ✓ | ✓ | 16/21 wellbeing services counties |
| Greece | 1 | ✓ |  | Min. of Health |
| Ireland | 1 | ✓ |  | HSE |
| Malta | 1 | ✓ |  | Min. of Health |
| Norway | 2 | ✓ | ✓ | 3/4 health regions |
| Slovenia | 1 | ✓ | ✓ | CRPD (national registry) |
| Sweden | 2 | ✓ |  | 19/21 regions |
| *Regional adoption* |  |  |  |  |
| Australia | 1 | ✓ | ✓ | 170 institutions<sup>*a*</sup> incl. 2 regional |
| Italy | 1 | ✓ | ✓ | Abruzzo (regional) + 1 institution |
| Netherlands | 3 | ✓ | ✓ | > 21 organisations |
| Spain | 2 | ✓ | ✓ | Catalonia (regional) + institutional |
| United Kingdom | 2 | ✓ | ✓ | > 27 organisations, mainly NHS trusts and health boards |
| *Institutional adoption* |  |  |  |  |
| China | 1 | ✓ | ✓ | 7 hospitals |
| Cyprus | 1 | ✓ |  | Public hospital network(30) |
| Germany | 2 | ✓ | ✓ | > 25 institutions |
| India | 1 | ✓ | ✓ | 2 institutions |
| Indonesia | 1 | ✓ |  | 1 hospital group (52 hospitals) |
| Lithuania | 1 | ✓ |  | 1 hospital |
| Luxembourg | 1 |  | ✓ | 1 institution |
| Nigeria | 1 | ✓ |  | 1 institution |
| Switzerland | 1 | ✓ |  | 2 hospitals |

<sup>*a*</sup> Single vendor’s aggregate. The 170 institutions were not individually named and could not be independently verified.

**Figure 1.**
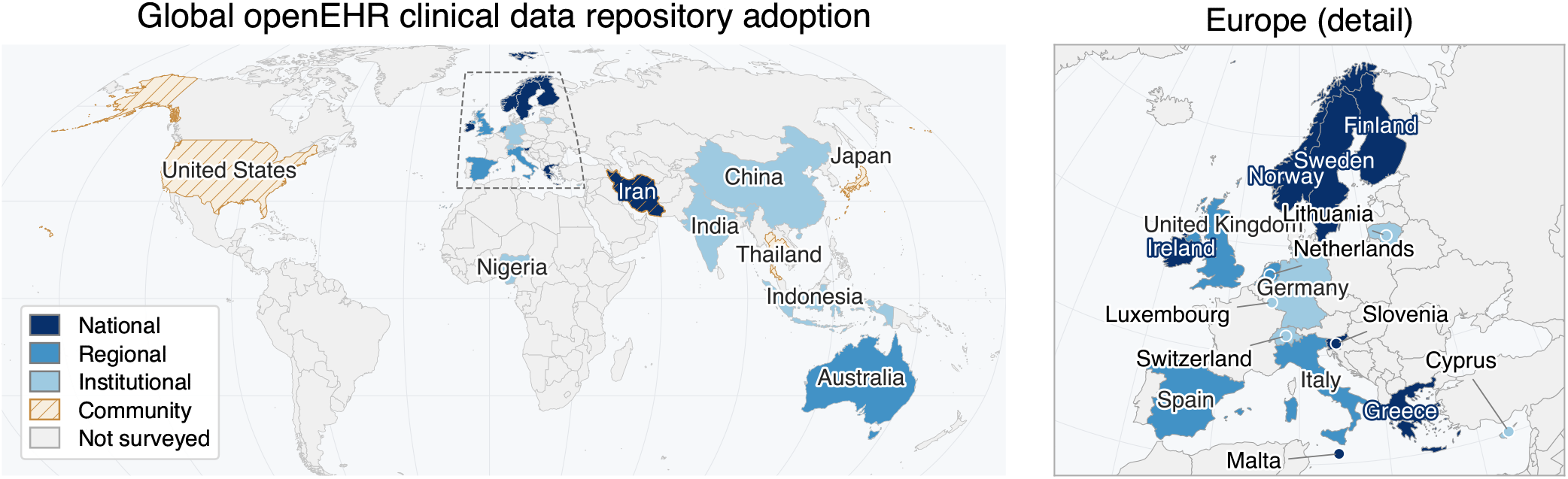
openEHR CDR adoption by country. Fill colour shows deployment scope (national = dark blue, regional = medium blue, institutional = light blue), with grey for countries covered by neither survey. Small countries carry a colour-coded dot. Diagonal hatching marks community-survey-only countries with no vendor response. Iran is national in scale but community-reported, shown as hatching over a national fill. The Australian entry is a single vendor’s unnamed aggregate of 170 institutions and is classified as regional. All figures are self-reported and unverified. The right panel shows European detail. N = 25 countries, 21 vendor-reported and 4 community-reported.

We classified each country’s adoption along two dimensions, scale (national, regional, or institutional) and adoption channel (Table 1). Individual hospital and hospital group deployments are noted descriptively where data permitted. In the vendor-market channel, health authorities procure an openEHR-based CDR from a commercial supplier. At national scale, vendor-market convergence produced high coverage. In Sweden, openEHR-based systems serve as the primary EHR across 19 of 21 health regions. In Norway, openEHR-based systems cover 3 of 4 regional health authorities, with the fourth operating a proprietary system. In Finland, an openEHR-based integrated health and social care system is in use across 16 of 21 wellbeing services counties. At regional and institutional levels, vendor-market adoption accounts for deployments across Australia (170 institutions, including two regional deployments, in the Northern Territory and Queensland (28)), the United Kingdom (several regional and institutional deployments), Germany (university hospitals and research institutions), the Netherlands (mental health, elderly care, disability care, and rehabilitation), Spain (Catalonia, regional), and several additional countries. Unlike the Nordic cases, the Australian figure is a single vendor’s unverified aggregate and the national coverage fraction is unknown, the regional classification follows from the two territory- and state-level deployments.

In the national health record channel, a government body has built or procured a national health record system whose technical foundation is openEHR. We define a national health record as a health-record system established or procured by a national government or ministry as country-wide health infrastructure, independent of how far its rollout has yet progressed. The system is governed and branded as national health infrastructure. openEHR is the architectural layer underneath and is not necessarily named in policy or governance documents. Slovenia (CRPD, Central Registry of Patient Data), Ireland (Health Service Executive), Malta (Ministry of Health), and Greece (Ministry of Health) each fall into this category. The community survey additionally reported Iran’s government-built national EHR system (SEPAS) as openEHR-based.

Wales reported deployments across multiple independently procuring regional health boards, alongside a national digital health coordination body. In Germany, individual hospitals, university hospitals, and hospital groups adopted openEHR. Reported institutions differ substantially in size, so entry counts understate reach. A single entry may denote one hospital or a group operating dozens, and the German entries include large hospital groups. Expanding them using publicly available hospital group sizes, the more than 25 reported German institutions correspond to approximately 165 hospitals, an estimated upper bound of approximately 9% of the 1,841 German hospitals recorded by Destatis in 2024(29). This figure carries considerable uncertainty, as it is unclear whether all hospitals within a reported group actively use the CDR.

Across reported deployments, primary (clinical) use predominated. Of all reported deployment entries, 61% supported primary use only and 12% supported both primary and secondary use, 26% reported secondary use only and one entry did not specify a use type. The national-scale vendor-market deployments covering substantial fractions of national health systems were almost exclusively primary-use oriented. In Germany, both primary and secondary use were reported. Secondary use was notably concentrated in university hospitals serving as research data platforms within the German Medical Informatics Initiative(31, 18), while primary use was reported across a range of individual hospitals and hospital groups. Finland was a notable exception, reporting secondary use alongside primary deployment at national scale. Fig. 2 shows vendor-reported regional health system coverage for the three countries with explicit region-level data, alongside countries where a government or ministry body adopted openEHR as national health record infrastructure.

**Figure 2.**
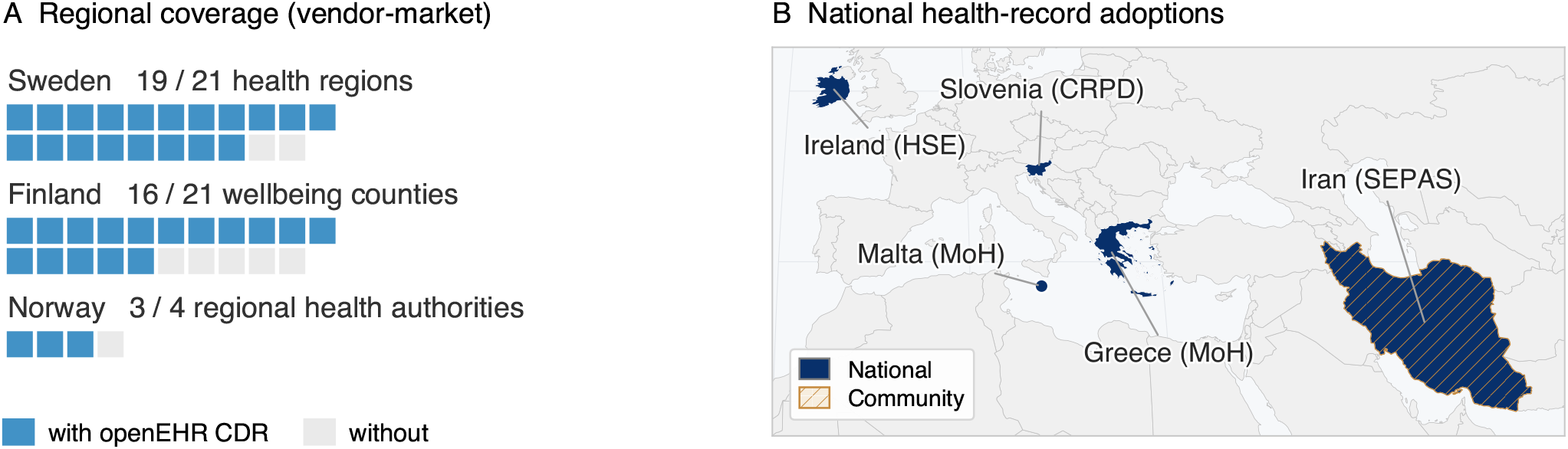
openEHR CDR adoption scale and channel. **(A)** Vendor-reported regional health system coverage for Sweden, Finland, and Norway, the only countries for which explicit numerator and denominator data were provided. **(B)** Countries where a government or ministry body adopted openEHR as the foundation of a national health record system. Abbreviations: CDR, clinical data repository; HSE, Health Service Executive; CRPD, Central Registry of Patient Data; MoH, Ministry of Health; SEPAS, Iranian national electronic health record system.

Free-text analysis identified four recurring themes, the role of the CDR, platform evolution, scaling patterns, and application domains. The first theme is the role the CDR plays, as a clinical system or as a data repository. In the national-scale deployments (Sweden, Finland, Norway), the openEHR CDR is embedded within or constitutes the clinical information system itself, meaning clinicians interact with it directly through the EHR interface and openEHR functions as the persistence layer underneath a complete clinical application. By contrast, some respondents described institutional-level deployments in which the openEHR CDR operates as a separate data repository alongside the existing hospital information system, with clinical data extracted from the primary HIS and stored in the openEHR CDR for secondary use.

The second theme is platform evolution. One respondent described a generational transition. Their first-generation CDR, integrated into an existing EHR system and deployed across the majority of Swedish health regions, supports primary use only. A second-generation standalone CDR designed for both primary and secondary use was under active rollout at the time of the survey, representing a platform upgrade within already-covered regions rather than new geographic expansion. Another respondent reported that their CDR functions as a complete EHR platform, blurring the outpatient and hospital distinction. The third theme is scaling. Two deployment patterns were described, single-instance deployments serving large organisational networks, for example 52 hospitals on one CDR instance, and incremental rollouts expanding from 1 to 10 to 25 hospitals in phases. The fourth theme is the range of application domains. Reported use cases extended beyond hospital CDRs to include elderly care, rehabilitation, mental healthcare, public health surveillance, and emergency data viewing, used as a backup when the primary hospital information system was unavailable.

## 4 DISCUSSION

This study provides the first vendor-based mapping of global openEHR CDR adoption. The deployment landscape is substantially broader and more mature than previously documented. A central pattern runs through the findings. openEHR adoption has been driven bottom-up by care providers, clinicians, and EHR vendors, not mandated top-down by governments or payers. Community survey respondents identified care providers (17/29) and innovation (17/29) as the primary adopting stakeholder and adoption driver respectively. Regulation and government grants accounted for only three responses. This is a standard chosen by practitioners without policy authorisation.

### 4.1 National-scale adoption without regulatory recognition

openEHR has achieved national-scale presence through two structurally distinct channels. In Sweden, Norway, and Finland, vendor market convergence has produced high regional coverage without a national programme. In Malta, Greece, Ireland, Iran, and Slovenia, national health authorities have built or procured health record systems whose technical foundation is openEHR. These systems are governed and branded as national EHR infrastructure rather than as openEHR deployments. These national-scale cases vary in evidentiary strength. The Nordic coverage figures and the Slovenian, Maltese, Greek(32), and Irish national records are corroborated by explicit coverage data, public policy documentation, or published accounts. The Iranian case rests on a community survey response, corroborated by a ministry-authored publication(33). Together, these channels document openEHR as a viable technical foundation for national health information infrastructure.

In no country covered by this survey did we identify openEHR named explicitly in binding national regulation. Slovenia comes closest, as national legislation governs the CRPD, which is built on openEHR, but the law names the system rather than the standard(34). Malta committed to openEHR through a formal national procurement tender, creating a contractual but not statutory obligation(35). Ireland’s Digital for Care 2024–2030 framework mentions openEHR alongside HL7 and FHIR as example standards, without mandate(36). Norway has established national archetype governance infrastructure through inter-regional collaboration, with archetypes maintained at a national library, but this is an organisational arrangement rather than a statutory requirement(37). Sweden, Finland, and Australia have no policy basis and adoption is entirely market-driven. The community survey reported openEHR activity in Iran, where the national EHR system (SEPAS) uses ISO 13606 for data transfer and openEHR for storage and modelling. The two standards are compatible given openEHR’s influence on ISO 13606. No Iranian regulation naming openEHR directly was identified.

The absence of regulatory recognition is consequential. National openEHR infrastructure built on procurement preference rather than legal mandate is vulnerable to reversal, and community survey respondents explicitly identified political direction changes as active risks. This gap is particularly relevant in the European context. The European Health Data Space (EHDS) mandates cross-border secondary use of health data and requires member states to establish interoperable national health data access bodies(38). openEHR-based CDRs are already operational at national or regional authority level across multiple EU member states, including Ireland, Malta, Greece, Slovenia, Finland, and Sweden, as well as Norway and the regions of Catalonia (Spain) and Abruzzo (Italy). These systems constitute precisely the semantically interoperable, persistent clinical data infrastructure that the EHDS envisions, yet this existing infrastructure is largely invisible to the EHDS implementation process(39).

The national patterns behind these figures differ by country. The concentration of vendor-market adoption in Nordic countries is consistent with the tradition of centralised, publicly funded eHealth strategies in the region(40), though a direct causal link between national eHealth policy and openEHR adoption cannot be established from our data. Australia is consistent with a market-driven adoption model. The UK’s more than 27 NHS organisations represent growing but not yet systemic adoption(41).

The broader regulatory environment is, however, moving in a favourable direction. International bodies increasingly frame interoperability and open, vendor-neutral standards as governance priorities. Recent OECD work on interoperability in healthcare(42) and the WHO draft reference architecture guidance for digital public infrastructure for health(43) both call for standards-based, interoperable national health-data infrastructure. The trajectory of a complementary standard is instructive. HL7 FHIR moved from specification to binding policy, mandated for health-data exchange in United States interoperability regulation, demonstrating that open standards can and do enter regulation. US federal policy has repeatedly shaped health information technology uptake in this way. Financial incentives under the HITECH Act drove large gains in hospital electronic health record adoption(44). openEHR is complementary to FHIR and occupies the persistence and modelling layer beneath it. Given the scale of adoption documented here, it is well positioned to move in a similar direction.

### 4.2 The secondary use gap

That 61% of deployments support primary use only, while just 12% support both primary and secondary use, reveals a structural gap despite openEHR’s architectural design for dual-purpose use(7). The community survey suggests that the barriers may be organisational, regulatory, and economic rather than primarily technical, consistent with the broader secondary use literature(24) and the “advanced use” divide in US hospitals(45, 46).

The community survey identifies a concrete contributor. In total, 21 of 29 respondents named *lack of openEHR-specific knowledge* as a primary barrier. This knowledge gap does not resolve once a national deployment exists. It persists and constrains what deployed systems can deliver. Countries with operational national CDRs still report insufficient openEHR expertise as an active barrier to unlocking secondary use.

Secondary-use deployments concentrate at German university hospitals serving the Medical Informatics Initiative. Unlocking secondary use in nationally deployed CDRs will likely require dedicated transformation infrastructure such as the Eos/OMOCL pipeline for openEHR-to-OMOP conversion(19) or the FHIRconnect bridge for openEHR-FHIR integration(21). More broadly, large language model-based ETL generation and semantic vocabulary mapping to the OMOP CDM may further lower this barrier(47).

### 4.3 openEHR and FHIR converge in practice

The community survey surfaced concurrent use of HL7 FHIR alongside openEHR as a dominant pattern, with FHIR used for data exchange and openEHR used for persistent clinical data storage. This is the complementary model proposed by Tsafnat et al.(8), reported as operational practice rather than theoretical possibility.

Two implications follow. First, the perceived competition between FHIR and openEHR is not reflected in deployment practice. Community survey respondents in the United Kingdom, the Netherlands, and Germany noted that decision-makers treat the two standards as mutually exclusive alternatives, whereas at the implementer level they are routinely deployed together. Framing a choice between them is a category error. Second, FHIR is the expected exchange standard for cross-border health data sharing under the EHDS, although it is not yet named in binding EU regulation. If openEHR deployments already operate alongside FHIR, the infrastructure documented here is partially integrated with the exchange layer the EHDS requires. Regulatory recognition is the missing component.

### 4.4 Vendor ecosystem and market dynamics

The eleven respondents span healthcare sectors well beyond hospital CDRs, including elderly care, mental healthcare, rehabilitation, public health surveillance, and emergency backup systems. This extends the product-focused characterisation of Delussu et al.(23) by documenting actual deployment contexts. In several countries, openEHR adoption has converged on a single supplier, making the standard and that supplier effectively synonymous in local markets. This substitutes one form of vendor dependency for another. The standard’s value proposition of vendor neutrality is not yet delivered by the market in these geographies. Broader supplier competition within the openEHR ecosystem would strengthen the standard’s core claim.

### 4.5 Limitations

Deployments are vendor self-reported and were not independently verified, although several of the larger ones are documented in the public domain. Respondents described systems they build or operate, so the reported figures carry a commercial interest and the main limitation is the absence of external corroboration. Some reported figures may reflect aspirational rather than fully operational states. For example, one respondent listed a hospital group comprising 16 hospitals as a single deployment entry while noting in a comment that the CDR was “currently in 1, soon rolling out to 10, and then 25 hospitals,” illustrating that the distinction between planned and operational deployments is not always clear in vendor-reported data.

Institution counts carry a second set of problems. Several responses listed hospital groups or associations as single entries without specifying the extent of deployment across constituent hospitals. For instance, one entry refers to a hospital operator comprising 79 hospitals and another to a hospital association with 22 member institutions, yet it could not be determined from the survey data whether the CDR is deployed across all constituent hospitals or only at the organisational level. Six of eleven respondents reported only aggregate figures (e.g., “170 institutions” or “16 regions”) without naming individual institutions, while the other five provided specific institution names. The actual number of institutions using openEHR CDRs cannot be precisely determined from the survey data. Institution counts are not directly comparable across respondents and should be interpreted as reported deployment entries rather than individual institution counts. One respondent contributed the majority of individually listed institutions, a reporting asymmetry reflecting response format rather than market share. Scope-based analysis mitigates this by focusing on country-level patterns.

Other limits concern what the survey reached at all. It did not capture deployment maturity or technical architecture, and deployment scope classification required interpretive judgement. The borderline cases were Australia and Cyprus, where the reported figures carried no coverage fraction. Assigning these two differently would move them by one tier without altering the overall pattern. In Jamaica, the vendor-reported national health record was a proof of concept that never went into production, consistent with both a published account(48) and the Jamaican community response. We therefore excluded it from the analysis. Delussu et al. identified 19 vendors(23), suggesting our 11 respondents represent approximately half the known market. The community survey further identified a government-built national openEHR deployment in Iran, reported by a single ministry-level respondent, and a direct follow-up with that respondent, who remains anonymous, reaffirmed the national-scale deployment. Independent verification was not possible given geopolitical access constraints. That case illustrates a systematic limitation of vendor-based surveys, namely that government-built, non-commercial deployments are invisible to vendor recruitment strategies.

Known non-response gaps include China (six vendors contacted, one responded) and Brazil (active openEHR community (49), no vendor response). Reasons for non-response were not collected. Since recruitment relied on openEHR Foundation networks and an English-language instrument, vendors whose openEHR activity is primarily domestically oriented were plausibly harder to reach and less inclined to respond. Peer-reviewed openEHR modelling work from China (50) further indicates domestic activity beyond what vendor recruitment captured. Institutional deployments are also known in Portugal (51), Saudi Arabia, and the United Arab Emirates, where no vendor responded. A city-wide openEHR deployment in Moscow was known to have been operational in the past, but no current information could be obtained due to the geopolitical situation. The survey captures a lower bound on global adoption. Vendor-based surveys are structurally blind to non-commercial deployments, and government-built, open-source, or internally developed systems are systematically underrepresented. The community survey respondents were self-selected through openEHR Foundation networks and their data are used mainly to contextualize vendor findings, for factual regulatory status, thematic patterns, and the identification of community-reported deployments. Both surveys were conducted at a single point in time and cannot capture the dynamic nature of CDR adoption.

## 5 CONCLUSION

This study provides the first mapping of global openEHR CDR adoption from vendor and community perspectives. openEHR has moved from a standards specification and research platform into production infrastructure across at least 21 countries. National-scale presence has been achieved through vendor-market convergence (Sweden, Norway, Finland) and national health record adoption (Malta, Greece, Ireland, Slovenia, and, by community report, Iran).

The surveys reveal a standard built from the ground up by clinicians, care providers, and vendors without government mandate. That bottom-up success has produced national-scale infrastructure that is structurally fragile. We identified no country that names openEHR in binding regulation. The infrastructure practitioners built can be reversed by the next procurement cycle.

Two priorities follow. First, the knowledge gap must be addressed. National-scale deployment has not eliminated the openEHR expertise deficit. It persists and constrains secondary use across all deployment tiers. Investment in education and training is as necessary as investment in technical infrastructure. Second, regulatory recognition must be pursued. Deployment evidence now exists across multiple countries and jurisdictions. The EHDS provides a concrete policy vehicle for formalising openEHR’s role in European health data infrastructure.

Future work should establish a longitudinal monitoring framework for openEHR adoption, extend survey methods to capture non-commercial and government-built deployments, and assess the impact of policy interventions on adoption patterns. Future surveys should distinguish the architectural role of the CDR, whether it operates as an integrated clinical information system, a standalone data persistence layer, or a dedicated secondary-use repository, as this distinction directly shapes the primary-secondary use balance.

## Supporting information

Supplementary Material: Survey instruments

## AUTHOR CONTRIBUTIONS

SK conceived and distributed the survey. SK and FME designed the study, analysed the data, and wrote the first draft. RE supervised the study and acquired funding. SK, FME, XM, NF, MM, and RE reviewed and revised the manuscript. All authors contributed to the article and approved the submitted version.

## ACKNOWLEDGMENTS

We thank all survey respondents for their participation.

## DATA AVAILABILITY STATEMENT

The survey instruments are provided in full as Supplementary Material. The aggregated country-level data supporting the conclusions of this article are available from the corresponding author on reasonable request. Individual vendor responses are not publicly available because respondents were assured of organisational anonymity.

