## Supplementary Material: Survey instruments for "Global Adoption of openEHR Clinical Data Repositories: A Vendor and Community Survey"

### Contents

- **S1** — Vendor survey instrument
- **S2** — Community survey instrument

Both instruments are reproduced below exactly as they were presented to respondents in Google Forms, including section structure, response options, and rating-scale anchors. Two items collected identifying information, namely country and organisation. These were used for country-level aggregation only, and individual responses are not reported. Both surveys opened with an information page and a consent item. Both are reproduced below, the consent item without a question number because it gates participation rather than collecting data. Some responses were clarified afterwards in writing or in conversation with the respondent. Those exchanges were unstructured and are therefore not reproduced here.

### S1 Vendor survey instrument

Survey title: Survey on Vendor Adoption of openEHR. 8 questions, preceded by the study information page and the consent item shown below.

#### Study information presented to respondents

**Survey Information** This survey is directed at vendors working with openEHR. The purpose is to examine where and how openEHR solutions are currently being implemented in healthcare institutions, including the extent of adoption and the main areas of application.

**Participation** Participation is entirely voluntary. You may skip any question or withdraw from the survey at any point without providing a reason.

**Use of Data** All responses will be analysed in aggregate to generate insights into the adoption of openEHR.

**Confidentiality** Information about reported clinical data repositories (CDRs) will be presented only in aggregated form and will not be linked to individual vendors. No personal or individual-level data will be collected or disclosed.

**Consent**

I have read the information above and consent to participate in this survey under the stated conditions.

**Response:** Single choice. (required)

- Yes
- No

### Contact information

**V1** Country

**Response:** Free text (short answer). (required)

**V2** Company

**Response:** Free text (short answer). (required)

### openEHR in institutions

**V3** In what Institutions is your CDR used ?

Answer format given in the form:
Institution, country
and/or
Nation/Region, country
as example:
Karloniska institutet, Sweden
Valencia, Spain

**Response:** Free text (long answer). (required)

**V4** In what Institution is your CDR used for BOTH primary (routine clinical use) and secondary use (research) ?

Answer format given in the form:
Institution, country
and/or
Nation/Region, country

**Response:** Free text (long answer).

**V5** In what Institution is your CDR used for primary use ONLY?

Answer format given in the form:
Institution, country
and/or
Nation/Region, country

**Response:** Free text (long answer).

**V6** In what Institution is your CDR used for secondary use ONLY ?

Answer format given in the form:
Institution, country
and/or
Nation/Region, country

**Response:** Free text (long answer).

**V7** Is the CDR used as an Outpatient Information Management System (GP, etc. )

Answer format given in the form:
Institution, country
and/or
Nation/Region, country

**Response:** Free text (long answer).

**V8** Is the CDR used as a Hospital Information System

Answer format given in the form:
Institution, country
and/or
Nation/Region, country

**Response:** Free text (long answer).

### S2 Community survey instrument

Survey title: Community Survey on openEHR Adoption and Use. 54 questions, preceded by the study information page and the consent item shown below.

#### Study information presented to respondents

**Study Information** This survey is directed at healthcare institutions and professionals with knowledge of openEHR adoption. The purpose is to understand where and how openEHR is being used, the extent of adoption, and the main areas of application.

**Participation** Participation is entirely voluntary. You may skip any question or stop the survey at any time without providing a reason.

**Use of Data** All responses will be analysed in aggregate to provide insights into national and regional adoption of openEHR. Results will be used exclusively for research and reporting.

**Confidentiality** The survey collects only organisational-level information (country and institution). No personal data are collected. Results will be presented in aggregate form, and no individual institution will be linked to specific responses.

**Consent**

I have read the study information above and consent to participate under the stated conditions.

**Response:** Single choice. (required)

- Yes
- No

#### Contact information (2 of 8)

Please provide your contact details.

**C1** Which country are you completing this survey for?

**Response:** Free text (short answer). (required)

**C2** Company/Institution

**Response:** Free text (short answer). (required)

#### General (3 of 8)

**C3** To what extent is openEHR used in your country for data persistence?

**Response:** Rating scale (1–5), from "not used at all" to "used as the main standard".

**C4** To what extend is openEHR used in your country for cross institutional interoperability?

**Response:** Rating scale (1–5), from "not used at all" to "used as the main standard".

**C5** To what extend is openEHR used in your country for clinical modeling?

**Response:** Rating scale (1–5), from "not used at all" to "used as the main standard".

**C6** What change in the adoption rate of openEHR do you expect in your country over the next years?

**Response:** Rating scale (1–5), from "strong decrease" to "strong increase".

#### Regulation (4 of 8)

**C7** Is there any regulation in your country that mandates the use of standards for electronic healthcare data exchange?

**Response:** Single choice.

- Yes
- No
- I don't know

**C8** If yes, does the regulation specifically mention openEHR ?

**Response:** Single choice.

- Yes, openEHR is mandated
- Yes, openEHR is advised
- No, openEHR is not mentioned
- I don't know

**C9** Is there any regulation in your country that mandates the use of standards in healthcare data persistence?

**Response:** Single choice.

- Yes
- No
- I don't know

**C10** If yes, does the regulation specifically mention openEHR ?

**Response:** Single choice.

- Yes, openEHR is mandated
- Yes, openEHR is advised
- No, openEHR is not mentioned
- I don't know

**C11** Is there a deadline for compliance included in the regulation ?

**Response:** Single choice.

- Yes
- No
- I don't know

**C12** If yes, are there fines imposed in case the regulation is not met before the deadline ?

**Response:** Single choice.

- Yes
- No
- I don't know

**C13** Are there any government funding programs available to support the adoption of openEHR ?

**Response:** Single choice.

- Yes
- No
- I don't know

**C14** Can you share more information about these regulations and funds, including names and links ?

**Response:** Free text (long answer).

**C15** Is there anything else you'd like to mention regarding health data in your country ?

**Response:** Free text (long answer).

#### National standards development (5 of 8)

**C16** Is there any national health organization in your country?

**Response:** Single choice.

- Yes
- No
- I don't know

**C17** Please provide a link to this national organization (or organizations), or leave blank if you don't know.

**Response:** Free text (long answer).

**C18** Are there any regional or national projects based on openEHR ?

**Response:** Single choice.

- Yes, widely used
- Yes, used in a limited set of use cases
- Yes, planned
- No
- I don't know

**C19** If yes, is it a national or regional health record with a central/federated openEHR CDR ?

**Response:** Single choice.

- Yes
- No
- I don't know

**C20** Please share the link to the published national or regional openEHR models, or leave blank if you don't know.

**Response:** Free text (long answer).

**C21** Are there other openEHR projects developed in your country for more specific use cases ?

**Response:** Single choice.

- Yes, many
- Yes, a few
- No
- I don't know

**C22** If yes, for which of the following use cases?

**Response:** Select all that apply.

- Referrals / Continuity of care
- Imaging
- Provider Directory

- Immunizations
- Clinical Registries
- Genomics
- Document Exchange
- Prescriptions / Pharmacy
- Consent
- Diagnostic Orders/Reports
- Patient Access
- Public Health Reporting
- Clinical Decision Support
- Clinical summaries
- Other

**C23** If yes, which clinical domains do they cover ?

**Response:** Select all that apply.

- Primary Care / Family Medicine
- Internal Medicine
- Pediatrics
- Obstetrics & Gynecology (Women's Health)
- Surgery
- Emergency Medicine
- Anesthesiology
- Radiology / Imaging
- Pathology / Laboratory Medicine
- Psychiatry / Behavioral Health
- Neurology
- Cardiology
- Oncology
- Orthopedics
- Endocrinology
- Gastroenterology
- Nephrology
- Pulmonology / Respiratory Medicine
- Dermatology
- Infectious Diseases
- Geriatrics
- Rehabilitation / Physical Medicine
- Public Health & Preventive Medicine
- PREMs/PROMs
- Social care

**C24** What proportion of the models you use in your openEHR projects come from the international CKM? (Leave blank if you don't know)

**Response:** Rating scale (1–5), from "None" to "All of it".

**C25** Is there anything else you want to share on national standard development in your country ?

**Response:** Free text (long answer).

### openEHR and other standards (6 of 8)

**C26** Does your organization use any of the following standards or technologies apart from openEHR?

**Response:** Select all that apply.

- FHIR for communication
- FHIR terminology services
- FHIR demographics
- FHIR storage
- Snomed-ct
- Loinc
- ICD9
- ICD10
- ICD11
- ATC
- RxNorm
- OMOP CDM
- CDISC
- RedCap
- I2B2

**C27** Does your organization use any of the following standards or technologies in combination with openEHR?

**Response:** Select all that apply.

- FHIR for communication
- FHIR terminology services
- FHIR demographics
- FHIR storage
- Snomed-ct
- Loinc
- ICD9
- ICD10
- ICD11
- ATC
- RxNorm
- OMOP CDM
- CDISC
- RedCap
- I2B2

#### openEHR implementation (7 of 8)

**C28** Which stakeholders are adopting openEHR in your country?

**Response:** Select all that apply.

- Care providers
- Payers / Insurers
- EHR system vendors
- Diagnostic system vendors, like Imaging / Lab
- App developers
- Clinical Registries
- Research institutions
- Other

**C29** What are the main drivers for openEHR adoption in your country ?

**Response:** Select all that apply.

- Regulation and grants
- Improving health outcomes
- Improving care workflows
- Innovation
- Other

**To what extent are the following openEHR components used in your country?**

**C30** Archetype modelling

**Response:** Rating scale (1–5), from "Not at all" to "Widely used".

**C31** AQL

**Response:** Rating scale (1–5), from "Not at all" to "Widely used".

**C32** Task planning

**Response:** Rating scale (1–5), from "Not at all" to "Widely used".

**C33** Folders

**Response:** Rating scale (1–5), from "Not at all" to "Widely used".

**C34** Smart on openEHR

**Response:** Rating scale (1–5), from "Not at all" to "Widely used".

**C35** openEHR demographics

**Response:** Rating scale (1–5), from "Not at all" to "Widely used".

**C36** Data federation on openEHR

**Response:** Rating scale (1–5), from "Not at all" to "Widely used".

**C37** OpenEHR used with/for AI

**Response:** Rating scale (1–5), from "Not at all" to "Widely used".

**C38** Are you aware of any successful openEHR use cases in your country ?

**Response:** Single choice.

- Yes
- No

**C39** If yes, what are the main achievements of these use cases?

**Response:** Select all that apply.

- Lowered cost
- Improved healthcare outcomes
- Improved access to information
- Improved care workflows
- Other

**C40** If other, please specify:

**Response:** Free text (long answer).

**C41** Please list the successful openEHR use cases you are aware of.

**Response:** Free text (long answer).

**C42** What are the biggest openEHR challenges in your country?

**Response:** Select all that apply.

- High investment cost
- Unclear benefits
- Unclear regulations
- Lack of openEHR knowledge
- Changes in political direction
- Other

**C43** If other, please specify:

**Response:** Free text (long answer).

**C44** Are there any implementation use cases in the future you are looking forward to?

**Response:** Free text (long answer).

**C45** To what extent are you using open source vs. proprietary openEHR software?

**Response:** Rating scale (1–5), from “Exclusively open source” to “Exclusively properitary”.

**C46** Is there anything else you want to share about the implementation of openEHR in your country ?

**Response:** Free text (long answer).

#### Final questions (8 of 8)

**C47** What were the major achievements in openEHR adoption in your country during the last year?

**Response:** Select all that apply.

- Establishment of an national standard organization
- Development of an national openEHR model
- New regulation that prescribes the use of standards in electronic health exchange
- Development of new openEHR standards for more specific use cases
- Launch of pilot projects with selected healthcare stakeholders
- Expanded adoption of openEHR accross the healthcare ecosystem
- Other

**C48** If other, please specify:

**Response:** Free text (long answer).

**C49** Regarding last year’s openEHR adoption, we have made ...

**Response:** Single choice.

- ... much less progress than we expected
- ... less progress than we expected
- ... quite the progress we expected
- ... more progress than expected
- ... far more progress than expected

**C50** How satisfied are you with the adoption rate of openEHR in your country ?

**Response:** Single choice.

- Very dissatisfied
- Dissatisfied
- Neutral
- Satisfied
- Very satisfied

**C51** What next steps do you expect in openEHR adoption in your country over the coming year?

**Response:** Select all that apply.

- Establishment of a national standard organization
- Development of a national openEHR model
- New regulation that prescribes the use of standards in electronic health data exchange
- Development of new openEHR standards for more specific use cases
- Launch of pilot projects with selected healthcare stakeholders
- Expanded adoption of openEHR accross the healthcare ecosystem
- Other

**C52** If other, please specify:

**Response:** Free text (long answer).

**C53** To what extent do you agree with the following statement?

Answer format given in the form:
"Within the next three years, I anticipate that our country will reap the benefits of openEHR adoption, resulting in significant cost savings, improved care coordination, and a stronger digital health ecosystem—paving the way for future healthcare innovation."

**Response:** Single choice.

- Strongly disagree
- Disagree
- Neither nor
- Agree
- Strongly agree

**C54** Besides the current respondents of this survey, who are the main openEHR stakeholders needed to obtain a complete overview of openEHR adoption in your country?

Answer format given in the form:
Please list relevant organizations or specific contacts (with contact details, if possible), so we can reach out to them for this or a future survey.

**Response:** Free text (long answer).
